# Individual-level Social Vulnerability and Cognitive Function Among Older Adults in the United States National Health and Nutrition Examination Survey

**DOI:** 10.64898/2026.07.02.26356711

**Authors:** Laura Arboleda-Merino, Erika Walker, Samuel D. Fansler, Spruha Joshi, Belinda Needham, Sung Kyun Park, Kelly M. Bakulski

**Affiliations:** Department of Epidemiology, School of Public Health, University of Michigan, Ann Arbor, MI, USA; Department of Biostatistics, School of Public Health, University of Michigan, Ann Arbor, MI, USA; Department of Environmental Health Sciences, School of Public Health, University of Michigan, Ann Arbor, MI, USA

**Keywords:** Aging, epidemiology, Socioeconomic factors, Digit Symbol Substitution Test, Health disparities

## Abstract

**Background:** Cognitive function is central to healthy aging and independent living. Its impairment burdens individuals and healthcare systems alike. Social vulnerability, shaped by demographic and socioeconomic factors, may be associated with cognitive health, but reliance on community-level assessments limits understanding of individual-level risk.

**Methods:** We analyzed cross-sectional data from 5,989 adults aged ≥60 years in the National Health and Nutrition Examination Survey (NHANES) 1999-2002 and 2011-2014 cycles to examine the association between an individual-level Social Vulnerability Index (iSVI) and cognitive function. We constructed the iSVI by summing six individual-level indicators: income, education, health insurance, employment, housing tenure, and food security. Cognitive performance was measured using the Digit Symbol Substitution Test (DSST). Survey-weighted linear regression estimated associations between iSVI and DSST scores, adjusting for age, sex, race/Hispanic origin, and survey cycle. Effect sizes were interpreted relative to the adjusted association between age and DSST scores.

**Results:** Higher iSVI was associated with poorer cognitive performance, with each one-unit increase corresponding to a 5.30-point lower DSST score (95% CI-5.71,-4.89). For comparison, each additional year of age was associated with a 1.08-point lower DSST score (95% CI-1.14,-1.03), showing that the DSST difference associated with a one-unit increase in iSVI was similar in magnitude to approximately five years of age-related difference in DSST performance.

**Conclusions:** Higher individual-level social vulnerability was associated with lower cognitive performance among older US adults. Individual-level vulnerability measures can help identify older adults at risk for poor cognitive outcomes who may benefit from targeted screening and intervention.

## 1. INTRODUCTION

Cognitive function is a cornerstone of healthy aging and a fundamental contributor to well-being in later life. It encompasses key domains such as memory, attention, language, and executive function, which collectively support autonomy, social engagement, and overall quality of life among older adults.[1,2] Age-related changes in cognitive performance range from mild cognitive impairment to progressive neurodegenerative conditions, such as dementia.[2] In the United States (US), cognitive impairment represents a substantial public health burden.[3,4] Approximately 22% of adults aged ≥65 years have mild cognitive impairment and 10% have dementia.[4] As the US population continues to age, the number of older adults affected by cognitive impairment is expected to increase considerably, presenting substantial challenges for individuals, families, caregivers, and healthcare systems.[5] Identifying vulnerability factors associated with poorer cognitive function may help inform strategies for earlier risk identification and intervention.[6–9]

Cognitive function is shaped by a complex interplay of genetic, biological, and environmental factors across the lifespan.[6] These factors do not operate in isolation, and are further shaped by social and structural contexts.[6,7,9] Social vulnerability refers to the degree to which individuals, communities, or societies are at risk of harm from stressors such as natural disasters, economic hardship, and health emergencies.[10–12] In research and public health practice, social vulnerability is used to identify groups at higher risk of adverse outcomes when exposed to these stressors due to social, economic, and structural disadvantages.[11] Greater social vulnerability may reflect barriers to prevention and care, which can compound biological risk and contribute to disproportionate burdens of cognitive impairment and dementia.[8,9,13] Several indices have been developed to measure social vulnerability. One such index is the Social Vulnerability Index, constructed by the Centers for Disease Control and Prevention and the Agency for Toxic Substances and Disease Registry (CDC/ATSDR SVI).[12] The CDC/ATSDR SVI uses census tract indicators of socioeconomic status, household composition, racial and ethnic minority status, housing, and transportation to identify at-risk communities.[12] Higher SVI scores often reflect structural disadvantage and neighborhood-level deprivation, both relevant to cognitive health through pathways involving chronic stress, healthcare access, environmental exposures, social resources, and opportunities for healthy aging.[8,12,13] As disaster management and public health planning tools, area-based vulnerability indices have proven valuable for identifying communities where resources and interventions may be most needed.[12]

However, area-based measures often fail to capture individual-level differences.[14–17] Studies consistently find low agreement between neighborhood-or census-derived socioeconomic measures with individual-level measures. In a Montreal study of school children, area-based socioeconomic indices classified children into the same socioeconomic quintile as their individual-level socioeconomic status only 28.7% of the time, with weak correlations between area-and individual-level measures.[14] Similarly, in a large population-based Ontario cohort, individual-level and area-level income quintiles matched for only 29% of respondents, with a Kappa statistic of 0.11, showing little agreement beyond chance.[15] Even when allowing classification within one adjacent quintile, agreement was only moderate.[15] Individuals within the same neighborhood or census tract may differ substantially in income, education, occupation, healthcare access, and other factors relevant to cognitive aging.[11,16] Relying on area-level measures as proxies for individual-level vulnerability may introduce misclassification and obscure individual-level pathways linking social disadvantage to cognitive outcomes.[14–17]

To address this gap, we propose an individual-level Social Vulnerability Index (iSVI) incorporating individual-specific sociodemographic characteristics, household conditions, healthcare access, and other key social features, to examine differences in cognitive function among older adults. Unlike traditional area-based indices, iSVI is designed to characterize personal circumstances that may shape cognitive aging, access to prevention and care, and exposure to social and health-related stressors.[7–9,16] Although area-level SVIs have been increasingly used to study health disparities, the role of individual-level social vulnerability in cognitive aging remains understudied. This study examines the association between iSVI and cognitive function among older adults in the US. By distinguishing individual-level vulnerability from neighborhood-level context, this work may help identify older adults at elevated risk for poor cognitive outcomes and inform tailored strategies for cognitive health promotion, early detection, and supportive care.

## 2. METHODS

### 2.1 Data and study population

The National Health and Nutrition Examination Survey (NHANES) is a series of ongoing cross-sectional surveys conducted by the CDC through the National Center for Health Statistics (NCHS), each representing an independent, nationally representative sample of the non-institutionalized civilian US population.[18] Since 1999, public-use datasets have been published in two-year cycles. We drew participants from four survey cycles (1999-2000, 2001-2002, 2011-2012, and 2013-2014) during which NHANES administered cognitive testing. All NHANES participants provided informed consent, and the University of Michigan Institutional Review Board approved this secondary data analysis (HUM00194918). All original datasets are publicly available through NCHS [19] and online repositories.[20]

### 2.2 Cognitive function measurement

Cognitive functioning was assessed in NHANES participants aged ≥60 years using the Digit Symbol Substitution Test (DSST), a performance module from the Wechsler Adult Intelligence Scale that evaluates processing speed, sustained attention, and working memory.[21] Participants were given two minutes to match digits 1–9 with corresponding symbols (e.g., arrows, squares), with each correct entry scored as one point (maximum: 133).[21,22] DSST scores were analyzed as a continuous variable in primary analyses. For supplemental descriptive analyses, we dichotomized DSST scores at the survey-weighted 25th percentile (≤38 in our sample), with scores in the lowest quartile classified as low cognitive performance and quartiles 2–4 classified as normal cognitive performance for comparison.[23]

### 2.3 Individual-level Social Vulnerability Index (iSVI) measurement

Social vulnerability was evaluated for each participant using an iSVI adapted from the CDC/ATSDR SVI framework (Figure 1).[12] To develop this index, we used publicly available NHANES questionnaire data to obtain information on socioeconomic conditions, household structure, and healthcare access. The iSVI included six components related to social vulnerability: income, educational attainment, health insurance, housing tenure, employment status, and food security. For income, educational attainment, health insurance, and employment status, we used vulnerability cut-offs informed by the CDC/ATSDR SVI framework.[12] For housing tenure and food security, which are not part of the original CDC/ATSDR SVI, cut-offs were determined based on NHANES questionnaire responses and guided by the literature on these social determinants of health.[11,24,25] Each participant was assigned a binary score of 0 (vulnerability absent) or 1 (vulnerability present) for each component of the iSVI. In cases where responses indicated partial vulnerability, an intermediate score of 0.5 was assigned.

**Figure 1.**
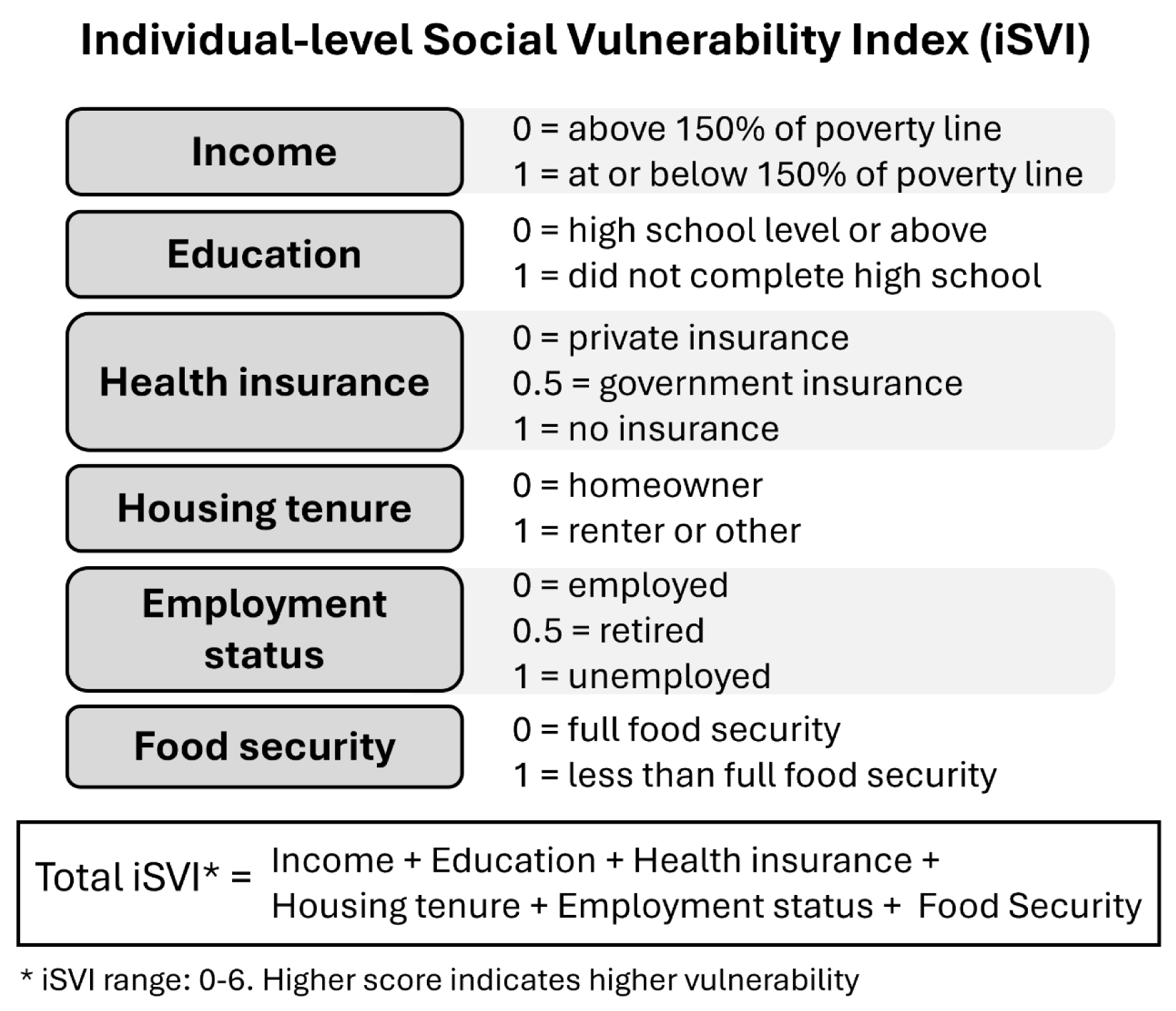
Components of the individual-level Social Vulnerability Index and its calculation.

Income was assessed using the poverty-income ratio, which compares family income with the federal poverty threshold for a given family size and survey year.[26] Poverty-income ratio was measured in NHANES as a continuous variable ranging from 0 (no income) to 5 (five times the poverty threshold or higher); values above 5 were recoded to protect confidentiality.[26] Participants received a score of 0 if their income was above 150% the poverty threshold (poverty-income ratio >1.5) and 1 if their income fell at or below that level.

Educational attainment was categorized as less than 9th grade, 9th to 11th grade (including 12th grade without a diploma), high school graduate or GED/equivalent, some college or associate’s degree, and college graduate or higher.[26] To align with the original CDC/ATSDR SVI threshold for educational attainment of “No high school diploma,” the lowest two categories were combined. Participants received a score of 0 if they completed high school or higher and 1 if they had not earned a high school diploma.

Health insurance status was recorded as private insurance, government-assisted coverage (including Medicaid, Medicare, military, or other public options), uninsured, or combinations of these types.[27] We collapse insurance status into three categories: private insurance, government-assisted insurance, and uninsured. Participants received a score of 0 for private insurance, 0.5 for government-assisted insurance, and 1 for no insurance. When multiple coverage types were reported, we prioritize private insurance for categorization.

Housing tenure was determined based on whether the respondent’s residence was owned, being purchased, rented, or occupied via another arrangement.[28] Participants were group as homeowners (owning or buying) or as renters/other arrangements and scored as 0 or 1, respectively.

Employment status was based on self-reported work during the previous week or, if unemployed, the reason for lack of work.[29] Participants were categorized as employed, retired, and not employed, with the latter group incorporating unemployed, family caregivers, students, those unable to work, disabled, or laid off. Participants received a score of 0 if they were employed, 0.5 if they were retired, and a 1 if they were not working; missing responses were labeled as such. Because retirement among older adults may reflect either normative life-course transitions or constrained labor-force exit, retired participants were assigned an intermediate vulnerability score.[29,30]

Finally, food security was measured based on participants’ self-report of household food conditions. Households were classified as fully food secure, marginally secure, food insecure without hunger, and food insecure with hunger.[31] Participants received a score of 0 for full food security and a 1 for any degree of food insecurity or marginal security.

All six components scores were summed to create an overall iSVI score ranging from 0 (lowest vulnerability) to 6 (highest vulnerability) (Figure 1). The iSVI enabled a comprehensive assessment of individual-level social vulnerability and allowed the study to capture the spectrum of disadvantages present among participants. We treated iSVI as both a continuous and a categorical variable. For categorical analyses, we compared participants in the highest iSVI quartile with those in quartiles 1–3 to evaluate differences associated with high individual-level social vulnerability.

### 2.4 Covariate measures

Primary models included age, sex, race/Hispanic origin, and survey cycle year as covariates to account for potential confounding. NHANES top codes age at 85 years in the 1999-2002 cycles and at 80 years in the 2011-2014 cycles. Sex was categorized as male or female. Race/Hispanic origin was self-reported. Mexican American and other Hispanic participants were combined into one Hispanic category. Non-Hispanic participants were classified as non-Hispanic White, non-Hispanic Black, or other race/ethnicity, including multiracial individuals.[26]

Secondary models additionally adjusted for health and behavior characteristics, including smoking status (never, former, or current), alcohol consumption (<1 or ≥1 drinks/day), and body mass index (BMI), which was calculated from measured height and weight and modeled continuously.[19,32–34] These variables were included in secondary models because they may represent mediating pathways through which social vulnerability influences cognitive performance.[16,35,36] Specifically, individual-level social vulnerability may shape smoking behavior, alcohol use, and BMI through chronic stress, socioeconomic constraints, reduced access to health-promoting resources, and barriers to preventive health care.[8,13,34,37] Therefore, adjustment for these factors may partially capture mechanisms linking social vulnerability to cognition, potentially attenuating the estimated total association.[16,35,36]

### 2.5 Statistical analysis

All analyses were performed in R version 4.5.0.[38] Participants aged ≥60 years with a valid DSST measure were included in the analytic sample, and a flow chart was used to visualize participant inclusion. Analysis accounted for the complex NHANES survey design by incorporating clustering, stratification, and sampling weights to produce nationally representative estimates.[39] Descriptive statistics compared characteristics of included versus excluded participants, with continuous variables summarized as means and standard deviations, and categorical variables presented as unweighted counts and percentages.

Within the analytic sample, missing data for iSVI components and covariates were imputed using multiple imputation by chained equations (MICE), generating five complete datasets.[40,41] The imputed variables are described in Supplemental Table 1. All models were run across each imputed dataset and pooled together using Rubin’s rules to create one set of estimates and measures of significance per model.[40]

Weighted descriptive statistics were calculated for included participants overall and stratified by iSVI category. The iSVI was categorized based on survey-weighted quartiles and collapsed into two groups: Low social vulnerability (quartiles 1-3) and high social vulnerability (quartile 4). In supplemental descriptive analyses, participant characteristics were also summarized by cognitive performance group: Low cognitive performance (quartile 1; DSST ≤38) and normal cognitive performance (quartiles 2-4; DSST >38). Group differences were evaluated using design-based Kruskal-Wallis tests for continuous variables and Rao-Scott adjusted Pearson chi-square tests for categorical variables. Variables contributing to the iSVI are highlighted in grey in the tables.

Within the analytic sample, we used histograms to visualize the distributions of DSST and iSVI scores, which allowed us to assess skewness, outliers, and other distributional features relevant for subsequent analysis. We also visualized associations between iSVI and DSST using survey-weighted scatterplots with linear trend lines, stratified by iSVI categories and weighted by sample size (Figure 2).

**Figure 2.**
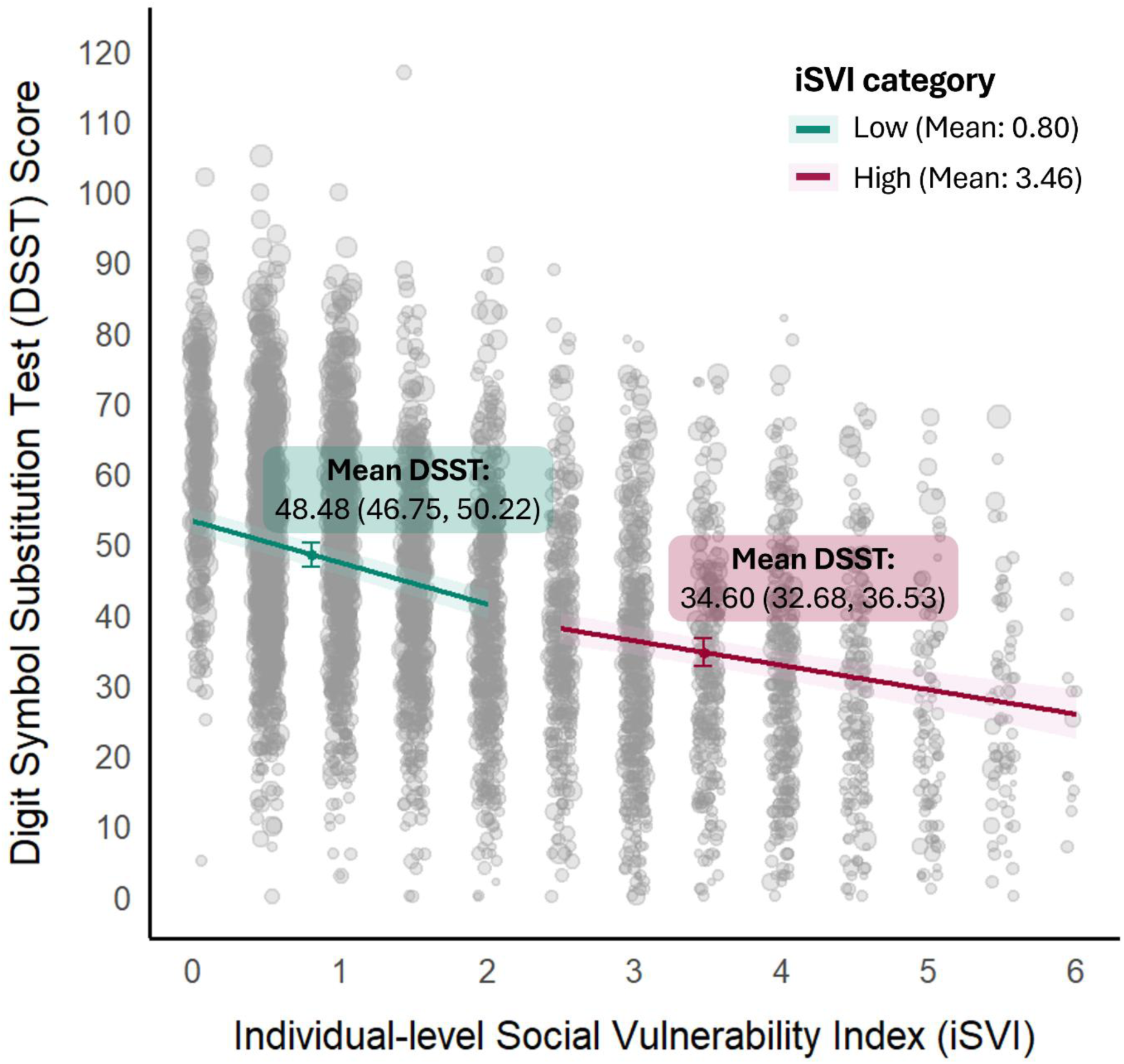
Adjusted association between individual-level Social Vulnerability Index and cognitive function measured using the Digit Symbol Substitution Test Score, stratified by individual-level Social Vulnerability Index levels in the US National Health and Nutrition Examination Survey. Non-imputed sample (N=5,272).

To evaluate the association between iSVI and cognitive function, we fit survey-weighted linear regression models. We first fit a model estimating the crude association, then fit a model adjusted for age, sex, race/Hispanic origin, and survey cycle. A secondary health and behavior model additionally adjusted for smoking status, alcohol consumption, and BMI.[32–34] Because these factors may lie on the causal pathway between iSVI and cognitive function, this model was not considered the primary adjustment model but was used to evaluate attenuation of the association after accounting for health behaviors and body composition. iSVI was first modeled continuously, with beta coefficients and 95% confidence intervals reported as the difference in DSST score per one-unit increase in iSVI. Next, we evaluated iSVI categorically, using low iSVI (quartiles 1-3) as the reference group, to facilitate comparisons across levels of individual-level social vulnerability. For comparison, we fit an additional model estimating the age-related difference in DSST scores, adjusted for sex, race/Hispanic origin, and NHANES cycle. From this model, we reported the difference in DSST score associated with each one-year increase in age. Statistical significance was defined as p < 0.05.

### 2.6 Sensitivity analyses

We conducted two sensitivity analyses. First, to examine whether the association between iSVI and DSST was driven primarily by any single iSVI component, we repeated the primary adjusted model six times, each time adding one component as an additional covariate: income, education, health insurance, housing tenure, employment and food security. This approach allowed us to assess whether adjustment for any individual component substantially attenuated the overall iSVI and DSST association, which would suggest that the observed association may be largely attributable to that specific domain of social vulnerability.

Second, we assessed potential effect modification by age, sex, and race/Hispanic origin by adding interaction terms between iSVI and each demographic covariate to the primary adjusted model. These analyses were motivated by evidence that the association between social vulnerability and cognition may vary across demographic groups because of differences in life-course exposure to social disadvantage, structural inequities, and cognitive aging trajectories.[7]

## 3. RESULTS

### 3.1 Participant characteristics

Among the 7,338 participants aged ≥60 years across the four selected NHANES cycles, 5,989 had complete DSST scores and were included in the analysis (Supplemental Figure 1). Excluded participants tended to be older, less likely to identify as Non-Hispanic White, had lower educational attainment, exhibited higher average iSVI scores, and reported slightly lower alcohol intake than those included (Supplemental Table 2). Included participants were on average 69.87 years old, 55.3% were female, and 81.0% identified as Non-Hispanic White (Table 1). Average DSST score was 49.61 (SD 17.73) and mean iSVI was 1.36 (SD 1.28) (Table 1; Supplemental Table 3). Within the analytic sample, 2,446 individuals were classified as having low cognitive performance (DSST ≤38; weighted 25th percentile; Supplemental Figure 2). Compared with participants in the comparison group (DSST >38; n = 3,543), participants with low cognitive performance were older, had higher iSVI scores, lower educational attainment, and were less likely to identify as Non-Hispanic White (Supplemental Table 4).

**Table 1.**
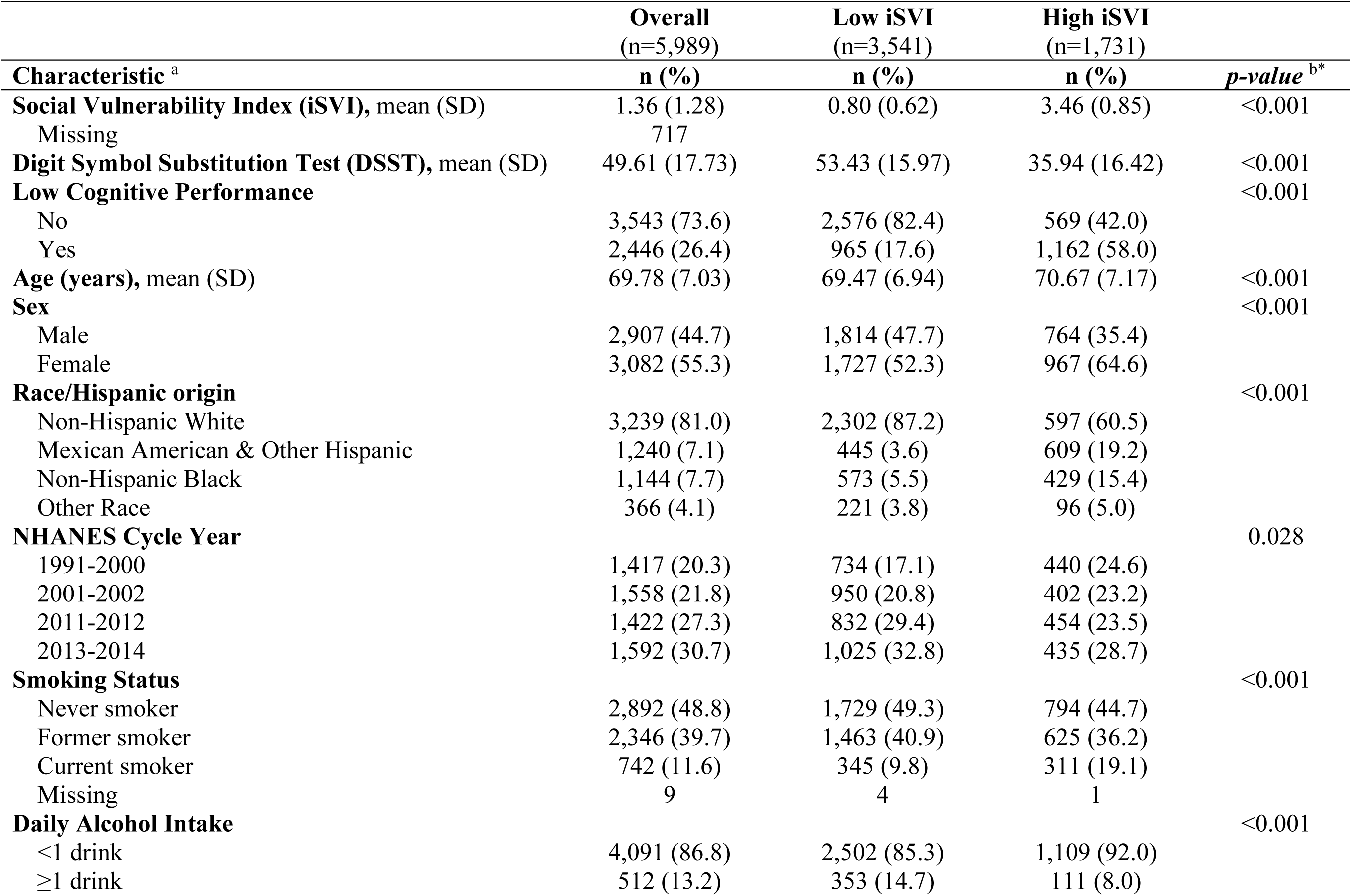
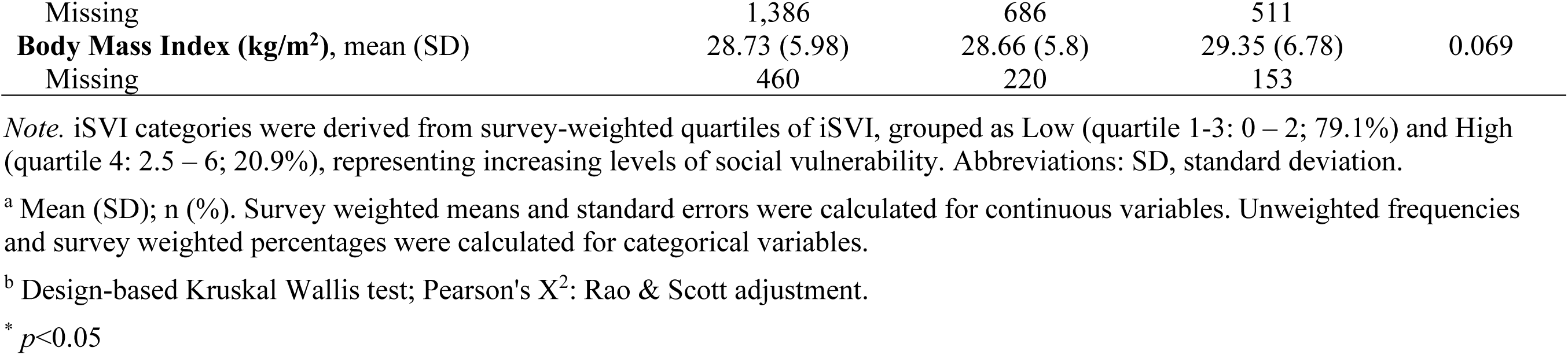
Weighted baseline characteristics of participants aged 60 years or older in the National Health and Nutrition Examination Survey, overall and stratified by individual-level Social Vulnerability Index categories.

|  | **Overall**<br>(n=5,989) | **Low iSVI**<br>(n=3,541) | **High iSVI**<br>(n=1,731) |  |
| --- | --- | --- | --- | --- |
| **Characteristic<sup>a</sup>** | **n (%)** | **n (%)** | **n (%)** | **p-value<sup>b\*</sup>** |
| **Social Vulnerability Index (iSVI), mean (SD)** | 1.36 (1.28) | 0.80 (0.62) | 3.46 (0.85) | <0.001 |
| Missing | 717 |  |  |  |
| **Digit Symbol Substitution Test (DSST), mean (SD)** | 49.61 (17.73) | 53.43 (15.97) | 35.94 (16.42) | <0.001 |
| **Low Cognitive Performance** |  |  |  | <0.001 |
| No | 3,543 (73.6) | 2,576 (82.4) | 569 (42.0) |  |
| Yes | 2,446 (26.4) | 965 (17.6) | 1,162 (58.0) |  |
| **Age (years), mean (SD)** | 69.78 (7.03) | 69.47 (6.94) | 70.67 (7.17) | <0.001 |
| **Sex** |  |  |  | <0.001 |
| Male | 2,907 (44.7) | 1,814 (47.7) | 764 (35.4) |  |
| Female | 3,082 (55.3) | 1,727 (52.3) | 967 (64.6) |  |
| **Race/Hispanic origin** |  |  |  | <0.001 |
| Non-Hispanic White | 3,239 (81.0) | 2,302 (87.2) | 597 (60.5) |  |
| Mexican American & Other Hispanic | 1,240 (7.1) | 445 (3.6) | 609 (19.2) |  |
| Non-Hispanic Black | 1,144 (7.7) | 573 (5.5) | 429 (15.4) |  |
| Other Race | 366 (4.1) | 221 (3.8) | 96 (5.0) |  |
| **NHANES Cycle Year** |  |  |  | 0.028 |
| 1991-2000 | 1,417 (20.3) | 734 (17.1) | 440 (24.6) |  |
| 2001-2002 | 1,558 (21.8) | 950 (20.8) | 402 (23.2) |  |
| 2011-2012 | 1,422 (27.3) | 832 (29.4) | 454 (23.5) |  |
| 2013-2014 | 1,592 (30.7) | 1,025 (32.8) | 435 (28.7) |  |
| **Smoking Status** |  |  |  | <0.001 |
| Never smoker | 2,892 (48.8) | 1,729 (49.3) | 794 (44.7) |  |
| Former smoker | 2,346 (39.7) | 1,463 (40.9) | 625 (36.2) |  |
| Current smoker | 742 (11.6) | 345 (9.8) | 311 (19.1) |  |
| Missing | 9 | 4 | 1 |  |
| **Daily Alcohol Intake** |  |  |  | <0.001 |
| <1 drink | 4,091 (86.8) | 2,502 (85.3) | 1,109 (92.0) |  |
| ≥1 drink | 512 (13.2) | 353 (14.7) | 111 (8.0) |  |
| Missing | 1,386 | 686 | 511 |  |
| **Body Mass Index (kg/m<sup>2</sup>), mean (SD)** | 28.73 (5.98) | 28.66 (5.8) | 29.35 (6.78) | 0.069 |
| Missing | 460 | 220 | 153 |  |

*Note.* iSVI categories were derived from survey-weighted quartiles of iSVI, grouped as Low (quartile 1-3: 0 – 2; 79.1%) and High (quartile 4: 2.5 – 6; 20.9%), representing increasing levels of social vulnerability. Abbreviations: SD, standard deviation.
<sup>a</sup> Mean (SD); n (%). Survey weighted means and standard errors were calculated for continuous variables. Unweighted frequencies and survey weighted percentages were calculated for categorical variables.
<sup>b</sup> Design-based Kruskal Wallis test; Pearson's X<sup>2</sup>: Rao & Scott adjustment.
\* $p < 0.05$

When iSVI was categorized (Table 1; Supplemental Figure 3), participants with high iSVI exhibited lower cognitive performance compared to those with low iSVI (mean DSST: 35.94, SD: 16.42 vs. 53.43; SD:15.97). The prevalence of low cognitive performance was also higher in the high iSVI group (58.0% vs. 17.6%). All characteristics, except for BMI, differed significantly across iSVI groups (p < 0.05), with individuals in the high iSVI group more likely to be female, older, Mexican American/Other Hispanic or Non-Hispanic Black, and less likely to be Non-Hispanic White (Table 1).

### 3.2 iSVI and cognitive function

Consistent with descriptive analyses, linear regression results showed a strong and persistent negative association between iSVI and DSST scores (Table 2). In the crude model, each one-unit increase in iSVI was associated with a 6.81-point lower DSST score (β =-6.81; 95% CI:-7.32,-6.31; p < 0.001), and participants in the high iSVI group had scores 17.69 points lower than those in the low iSVI group (β = - 17.69; 95% CI:-19.34,-16.04; p < 0.001).

**Table 2.** Associations between individual-level Social Vulnerability Index and Digit Symbol Substitution Test of participants aged 60 years or older in the National Health and Nutrition Examination Survey.

| Models | Continuous of iSVI |  | Categories of iSVI |  |  |
| --- | --- | --- | --- | --- | --- |
| | Per one unit increase<br>$\beta$ (95% CI) | *p*-value | Low | High<br>$\beta$ (95% CI) | *p*-value |
| Crude | **-6.81**<br>(-7.32, -6.31) | <0.001 | Ref | **-17.69**<br>(-19.34, -16.04) | <0.001 |
| Primary <sup>a</sup> | **-5.30**<br>(-5.71, -4.89) | <0.001 | Ref | **-13.49**<br>(-14.86, -12.13) | <0.001 |
| Health and behavior <sup>b</sup> | **-5.11**<br>(-5.53, -4.70) | <0.001 | Ref | **-13.00**<br>(-14.48, -11.53) | <0.001 |

*Note.* iSVI categories were derived from survey-weighted quartiles of iSVI, grouped as Low (quartile 1-3: 0 – 2; 79.1%) and High (quartile 4: 2.5 – 6; 20.9%), representing increasing levels of social vulnerability. Abbreviations: CI, confidence interval; iSVI, individual-level Social Vulnerability Index; Ref, Reference category.
<sup>a</sup> Adjusted for age, sex, race/Hispanic origin, survey cycle
<sup>b</sup> Adjusted for age, sex, race/Hispanic origin, survey cycle, smoking status, alcohol consumption, and body mass index.
\* $p < 0.05$

In the primary model, adjusted for age, sex, race/Hispanic origin, and NHANES cycle, higher iSVI remained significantly associated with lower DSST scores (Table 2). Each one-unit increase in iSVI was associated with a 5.30-point lower DSST score (β =-5.30; 95% CI:-5.71,-4.89; *p* < 0.001), and participants in the high iSVI group scored 13.49 points lower on the DSST than those in the low iSVI group (β =-13.49; 95% CI:-14.86,-12.13; *p* < 0.001). For comparison, a one-year increase in age was associated with a 1.08-point lower DSST score (95% CI:-1.14,-1.03) when adjusted for sex, NHANES cycle year, and race/Hispanic origin.

This association was slightly attenuated but remained statistically significant after additional adjustment for smoking status, alcohol consumption, and BMI (Table 2). Each one-unit increase in iSVI was associated with a 5.11-point lower DSST score (β =-5.11; 95% CI:-5.53,-4.70; *p* < 0.001), and high iSVI was associated with a 13.00-point lower DSST score compared with Low iSVI (β =-13.00; 95% CI:-14.48,-11.53; *p* < 0.001).

### 3.3 Sensitivity results

In sensitivity analyses evaluating whether the association between overall iSVI and DSST performance was driven by any single iSVI component, overall iSVI remained inversely associated with DSST scores after additional adjustment for each component individually (Table 3). When income, education, health insurance, housing tenure, employment, and food security were added separately to the primary adjusted model, the association remained generally consistent. Although adjustment for income and education modestly attenuated the magnitude of the association, higher overall iSVI continued to be associated with poorer cognitive performance across models. These findings suggest that the observed association between iSVI and DSST scores was not fully explained by any single component of iSVI.

**Table 3.** Sensitivity analyses of the association between individual-level Social Vulnerability Index and Digit Symbol Substitution Test of participants aged 60 years or older in the National Health and Nutrition Examination Survey.

| Models <sup>a</sup> | Continuous of iSVI |  | Categories of iSVI |  |  |
| --- | --- | --- | --- | --- | --- |
| | Per one unit increase<br>$\beta$ (95% CI) | *p-value* | Low | High<br>$\beta$ (95% CI) | *p-value* |
| Primary | **-5.30**<br>(-5.71, -4.89) | <0.001 | Ref | **-13.49**<br>(-14.86, -12.13) | <0.001 |
| Primary + Income | **-4.13**<br>(-4.64, -3.62) | <0.001 | Ref | **-8.72**<br>(-10.09, -7.35) | <0.001 |
| Primary + Education | **-3.56**<br>(-4.03, -3.08) | <0.001 | Ref | **-7.98**<br>(-9.49, -6.47) | <0.001 |
| Primary + Health insurance | **-5.54**<br>(-6.02, -5.05) | <0.001 | Ref | **-12.38**<br>(-13.83, -10.93) | <0.001 |
| Primary + Housing tenure | **-6.04**<br>(-6.51, -5.56) | <0.001 | Ref | **-13.52**<br>(-15.02, -12.02) | <0.001 |
| Primary + Employment | **-5.22**<br>(-5.67, -4.77) | <0.001 | Ref | **-12.21**<br>(-13.58, -10.84) | <0.001 |
| Primary + Food security | **-5.69**<br>(-6.16, -5.22) | <0.001 | Ref | **-12.65**<br>(-14.14, -11.15) | <0.001 |

*Note.* iSVI categories were derived from survey-weighted quartiles of iSVI, grouped as Low (quartile 1-3: 0 – 2; 79.1%) and High (quartile 4: 2.5 – 6; 20.9%), representing increasing levels of social vulnerability. Abbreviations: CI, confidence interval; iSVI, individual-level Social Vulnerability Index; Ref, Reference category.
<sup>a</sup> All models were adjusted for age, sex, race/Hispanic origin, NHANES cycle. Overall iSVI remained the primary exposure in all models, with each individual iSVI component added separately as an additional covariate to evaluate whether any single component explained the observed association.
\* $p < 0.05$

In effect modification analyses, we found little evidence that the association between iSVI and DSST scores varied by sex or race/Hispanic origin (Supplemental Table 5). Interaction terms for these characteristics were not statistically significant, and the inverse association between iSVI and DSST was generally consistent across subgroups. We observed some evidence of effect modification by age when iSVI was modeled categorically, with the association appearing stronger among participants younger than 70 years; however, this pattern was not observed when iSVI was modeled continuously. Overall, these findings suggest that the association between higher iSVI and lower DSST scores was robust, not driven by a single iSVI component, and largely consistent across demographic subgroups.

## 4. DISCUSSION

We evaluated the association between individual-level social vulnerability and cognitive function in a nationally representative sample of 5,989 US adults aged ≥60 years across four NHANES cycles. Higher iSVI scores, reflecting greater individual-level social vulnerability, were associated with poorer cognitive function. This association persisted in survey-weighted models adjusted for age, sex, race/Hispanic origin, and NHANES cycle, and results were consistent when iSVI was modeled as both a continuous and categorical measure. The cross-sectional DSST difference associated with each one-unit increase in iSVI was similar in magnitude to the adjusted DSST difference associated with nearly five additional years of age. The difference comparing participants with high versus low iSVI corresponded to more than a decade of age-related difference. These findings underscore substantial cognitive differences by individual-level social vulnerability among older US adults. Sensitivity analyses further supported the robustness of these findings, suggesting that the association was not fully explained by any single iSVI component and was generally consistent across demographic groups.

Our findings align with a growing body of literature linking greater social vulnerability and social disadvantage to poorer cognitive outcomes in older adults. Social vulnerability refers to an individual’s relative susceptibility (or resilience) to adverse health and functional outcomes in response to changes in social circumstances and environmental stressors, reflecting the cumulative impact of social deficits and supports.[11] It is commonly operationalized using composite indices that aggregate multiple social indicators into a single measure intended to capture this complexity and predict clinically meaningful outcomes. Prior studies using neighborhood and community-based measures, such as the Area Deprivation Index (ADI) [42–44], and the CDC/ATSDR SVI [12,45,46], as well as a few individual-level vulnerability indices [47,48] have generally observed a similar trend: greater social disadvantage is associated with lower cognitive performance, faster cognitive decline, and higher risk of cognitive impairment and dementia. For example, in the population-based Mayo Clinic Study of Aging, higher neighborhood socioeconomic disadvantage measured by the ADI was associated with greater odds of mild cognitive impairment (OR 1.13; 95% CI 1.08 - 1.19 per decile increase in national ADI rank).[42] Similarly, in a longitudinal study of cognitively unimpaired middle-to older-aged adults (mean age 59), living in the 20% most disadvantaged neighborhoods (by ADI) was associated with greater cortical thinning in Alzheimer’s disease signature regions and cognitive decline on a preclinical Alzheimer’s disease cognitive composite.[44]

Among the few studies that have directly evaluated the associations between SVI and cognition, findings have been broadly consistent with ours, though most relied on area-level measures. A study in 780 older adults in Chicago found that area-level SVI was associated with baseline global cognitive function in Black participants, though not with global cognitive change over time.[45] In a population-based cohort of 6,781 adults aged ≥65 years, higher neighborhood social vulnerability, measured by the CDC/ATSDR SVI, was associated with 18%-25% faster global cognitive decline, and roughly twofold higher odds of incident clinically diagnosed Alzheimer disease.[46] Two studies incorporated individual-level social vulnerability data. In a Canadian cohort of 2,468 participants aged ≥70 years, high social vulnerability was associated with 36% increased odds of experiencing cognitive decline compared to low social vulnerability.[47] In the prospective Honolulu-Asia Aging Study of 3,845 Japanese American men aged 71 to 93, higher baseline social vulnerability was associated with greater subsequent decline in global cognitive screening scores.[48]

Although direct comparison across studies is limited by differences in SVI measures, cognitive outcomes, and study design, the magnitude of our findings was substantial: high versus low iSVI was associated with a 13.49-point lower DSST score, similar in magnitude to the adjusted DSST difference associated with more than 12 additional years of age. Our findings extend prior work by showing that individual-level social vulnerability is strongly associated with lower cognitive performance in a nationally representative sample of older US adults.

By using an individual-level composite index rather than examining single socioeconomic predictors or relying solely on area-based measures, our study addresses a key gap by capturing the cumulative burden of multiple, co-occurring vulnerability factors at the individual level.[11,14,15,47,48] This approach addresses two important limitations of prior work. First, although individual predictors such as income, education, employment, insurance status, or food insecurity have each been linked to cognitive outcomes,[8,9,13,47] examining these factors separately may not fully capture the ways social risks cluster within the same person or how cumulative vulnerability may be associated with cognitive health. Social risks such as low income, lower educational attainment, lack of health insurance, housing instability, unemployment, and food insecurity often co-occur and may be jointly associated with cognitive function in ways not fully represented by any single factor.[8,13,47] The iSVI reflects this accumulation of social risk rather than the independent contribution of one socioeconomic characteristic.

Second, area-level vulnerability indices such as the ADI and the CDC/ATSDR SVI, are valuable for characterizing neighborhood conditions and guiding public health planning. However, they are not designed to measure social vulnerability at the level where many clinical and behavioral risks operate, namely the individual. Individuals within the same neighborhood or census tract may differ substantially in income, education, health care access, and other factors relevant to cognitive aging.[11,16] Assigning the same area-level score to all residents may obscure within-neighborhood heterogeneity, introduce ecological misclassification, and attenuate effect estimates if area-level measures do not accurately reflect individual circumstances.[14–17,49] Additionally, using area-level data to guide patient-level interventions is prone to the ecological fallacy, meaning incorrect assumptions about individuals based on the profile of their group.[16,49] An individual-level index provides a complementary approach by directly capturing personal circumstances across multiple social domains, improving precision for epidemiologic analyses and offering a more appropriate foundation for identifying older adults who may benefit from additional screening, outreach, or supportive services.

Although the composite index does not identify one specific vulnerability factor for intervention, it may help identify older adults at higher risk for poorer cognitive function who could benefit from targeted screening, resource referral, and linkage to social supports. In this way, the iSVI may be most useful as a risk-stratification tool rather than as a single-domain intervention guide. For example, older adults with high iSVI scores could be prioritized for cognitive screening, care coordination, social work referral, or connection to community-based resources. Potential interventions could include connecting high-risk individuals with food assistance, insurance navigation, housing resources, income-support programs, employment or disability services, and cognitive health services. Because the iSVI components are individually measurable, the index could first identify individuals at elevated risk, after which clinicians, public health practitioners, or community organizations could evaluate which specific needs are most actionable for that person. Together, these findings highlight individual-level social vulnerability as an important marker of cognitive function in older adults and a potential tool for identifying populations at elevated risk for cognitive disparities.

Several limitations should be considered when interpreting our findings. First, the cross-sectional design prevents causal inference, and prospective studies are needed to characterize trajectories of cognitive decline and evaluate whether social vulnerability precedes cognitive changes. Residual confounding is also possible, particularly from unmeasured or imperfectly measured factors such as early-life socioeconomic conditions, education quality, lifetime occupation and wealth, depression, and other health or social characteristics that shape both vulnerability and cognition. Selection bias is also possible because participants excluded due to missing DSST scores tended to be older, had higher iSVI scores, and had lower educational attainment than those included in the analytic sample. If excluded participants also had poorer cognitive performance, this exclusion may have led to underrepresentation of the most socially vulnerable older adults with low cognitive performance, potentially attenuating the observed association between iSVI and DSST performance. In addition, our reliance on the DSST, while a robust measure of processing speed, sustained attention, and working memory,[21] does not capture the full range of cognitive domains, and future research should incorporate broader cognitive batteries, neuroimaging, and/or adjudicated clinical outcomes to provide a more comprehensive assessment.

Finally, although an iSVI may better approximate personal circumstances than an area-level index, it still cannot fully capture the underlying construct we aim to measure. The concept of social vulnerability reflects an individual’s relative vulnerability or resilience to disruptions in their environment, social circumstances, health, or functional well-being, and older adults’ social circumstances are complex and multidimensional, with social vulnerability likely reflecting both individual and neighborhood-level factors.[11,47,50] In addition, any index is shaped by measurement and construction decisions, including which social domains are included, how these components are coded, and how they are weighted, and the iSVI derived from NHANES represents a cross-sectional snapshot that may not capture life course timing, duration, or changes in vulnerability over time. A true global measure of social vulnerability would need to be broad enough to capture a rich description of social deficits, practical to measure in both population and clinical settings, responsive to meaningful change over time, and predictive of important health outcomes. Thus, the iSVI, like any composite index, should be viewed as a practical proxy rather than a complete representation of this ideal.[50]

Although our limitations warrant caution, the study also has several strengths. First, we used a nationally representative sample of nearly 6,000 older adults from four NHANES cycles with diverse racial, ethnic, and socioeconomic backgrounds. Our large sample size and weighted data allow us to reflect the diversity of the US older adult population, enhancing generalizability. Second, prior research using area-based measures of vulnerability often overlooks individual variation and does not fully capture individual-level social risk.[16,42,43] By applying a detailed, individual-level composite index, we capture how multiple social determinants of health accumulate and interact within the same person’s lived context, rather than treating each factor independently. This approach provides a more faithful representation of real-world vulnerability because risks often compound, and cumulative vulnerability may have greater implications for cognitive health than any single social risk factor alone.[11,47,48] In contrast, one-variable-at-a-time analyses, and especially neighborhood or area-based indices, can miss key patterns such as clustering of disadvantages, effect modification, and threshold effects where vulnerability escalates once multiple stressors accumulate. While neighborhood-level vulnerability remains an important contextual factor, the public-use NHANES data does not include the geocoded residential information needed to link participants to measures such as CDC/ATSDR SVI. Thus, although we could not assess whether neighborhood-level vulnerability confounded or modified the observed associations, our use of an individual-level index allowed us to capture within-person accumulation of social risks that area-based measures alone may obscure. Third, sensitivity analyses, including additional adjustment for individual iSVI components, health and lifestyle covariates, and tests of effect modification by age, sex, and race/Hispanic origin, supported the robustness of our results. By evaluating potential sources of bias, including confounding and selection bias, and leveraging rigorous exposure and outcome assessment alongside survey-weighted analytic methods, we provide well supported estimates of the association between social vulnerability and cognitive function. Together, these strengths improve our ability to identify older adults who may be disproportionately affected and to inform public health efforts aimed at reducing cognitive disparities.

## 5. CONCLUSION

In a nationally representative sample of US adults aged ≥60 years, higher individual-level social vulnerability was strongly and consistently associated with lower DSST performance, including when iSVI was modeled continuously and categorically. Together with prior neighborhood-based studies, these findings support social vulnerability as an important factor linked to cognitive health in later life and suggest that measuring vulnerability at the individual level may better identify older adults who experience disproportionate cognitive disadvantage. From a public health perspective, incorporating individual-level social vulnerability into aging research and population surveillance may help identify individuals at greatest risk and inform more targeted prevention, outreach, and supportive services aimed at reducing cognitive disparities. Future longitudinal studies should evaluate whether changes in social vulnerability precede cognitive decline, identify the most influential vulnerability domains and potential thresholds, and test whether interventions that address modifiable social risks or strengthen social supports can slow cognitive decline or narrow disparities.

## Ethics statement

Data collection for NHANES was approved by the NCHS Research Ethics Review Board (ERB). All NHANES participants provided informed consent, and the University of Michigan Institutional Review Board approved this secondary data analysis (HUM00194918).

## Author Contribution Declaration

Laura Arboleda-Merino: Conceptualization, Data curation, Formal analysis, Methodology, Software, Visualization, Writing - original draft, Writing - review & editing,

Erika Walker: Data curation, Validation, Writing - review & editing

Samuel D. Fansler: Conceptualization, Methodology, Writing - review & editing Spruha Joshi: Writing - review & editing

Belinda Needham: Writing - review & editing

Sung Kyun Park: Conceptualization, Funding acquisition, Methodology, Project administration, Supervision, Writing - review & editing

Kelly M. Bakulski: Conceptualization, Funding acquisition, Methodology, Project administration, Supervision, Writing - review & editing

## Data availability statement

All data are publicly available through the US National Center for Health Statistics. Code to produce these analyses and graphics is available through GitHub (https://github.com/bakulskilab/NHANES_iSVI_cognition).

## Acknowledgments

We thank the participants and staff of the National Health and Nutrition Examination Survey.

## GLOSSARY

ADI: Area Deprivation Index
ATSDR: Agency for Toxic Substances and Disease Registry
CDC: Centers for Disease Control and Prevention
CI: Confidence Interval
DSST: Digit Symbol Substitution Test
iSVI: Individual-level Social Vulnerability Index
NHANES: National Health and Nutrition Examination Survey
SVI: Social Vulnerability Index
US: United States

## REFERENCES

[1] Mitchell AJ, Kemp S, Benito-León J, et al. The influence of cognitive impairment on health-related quality of life in neurological disease. Acta Neuropsychiatrica 2010;22:2–13. 10.1111/j.1601-5215.2009.00439.x.

[2] Murman DL. The Impact of Age on Cognition. Semin Hear 2015;36:111–21. 10.1055/s-0035-1555115.

[3] Fang M, Hu J, Weiss J, et al. Lifetime risk and projected burden of dementia. Nat Med 2025;31:772–6. 10.1038/s41591-024-03340-9.

[4] Manly JJ, Jones RN, Langa KM, et al. Estimating the Prevalence of Dementia and Mild Cognitive Impairment in the US: The 2016 Health and Retirement Study Harmonized Cognitive Assessment Protocol Project. JAMA Neurol 2022;79:1242–9. 10.1001/jamaneurol.2022.3543.

[5] Khan HTA, Addo KM, Findlay H. Public Health Challenges and Responses to the Growing Ageing Populations. Public Health Challenges 2024;3:e213. 10.1002/puh2.213.

[6] Langa KM. Cognitive Aging, Dementia, and the Future of an Aging Population. Future Directions for the Demography of Aging: Proceedings of a Workshop, National Academies Press (US); 2018.

[7] Yang YC, Walsh CE, Shartle K, et al. An Early and Unequal Decline: Life Course Trajectories of Cognitive Aging in the United States. J Aging Health 2024;36:230–45. 10.1177/08982643231184593.

[8] Majoka MA, Schimming C. Effect of Social Determinants of Health on Cognition and Risk of Alzheimer Disease and Related Dementias. Clinical Therapeutics 2021;43:922–9. 10.1016/j.clinthera.2021.05.005.

[9] Trani J-F, Zhu Y, Walker AIB, et al. Association Between Structural and Social Determinants of Health and Cognitive Functioning Among African Americans: the ARCHES Cohort. J Racial and Ethnic Health Disparities 2025. 10.1007/s40615-025-02730-0.

[10] Cutter SL, Boruff BJ, Shirley WL. Social Vulnerability to Environmental Hazards. Social Science Quarterly 2003;84:242–61. 10.1111/1540-6237.8402002.

[11] Mah JC, Penwarden JL, Pott H, et al. Social vulnerability indices: a scoping review. BMC Public Health 2023;23:1253. 10.1186/s12889-023-16097-6.

[12] CDC/ATSDR Social Vulnerability Index (SVI) 2024. https://www.atsdr.cdc.gov/placeandhealth/svi/index.html (accessed April 25, 2024).

[13] Adkins-Jackson PB, George KM, Besser LM, et al. The structural and social determinants of Alzheimer’s disease related dementias. Alzheimer’s & Dementia 2023;19:3171–85. 10.1002/alz.13027.

[14] Demissie K, Hanley JA, Menzies D, et al. Agreement in measuring socio-economic status: area-based versus individual measures. Chronic Dis Can 2000;21:1–7.

[15] Buajitti E, Chiodo S, Rosella LC. Agreement between area-and individual-level income measures in a population-based cohort: Implications for population health research. SSM Popul Health 2020;10:100553. 10.1016/j.ssmph.2020.100553.

[16] Gottlieb LM, Francis DE, Beck AF. Uses and Misuses of Patient-and Neighborhood-level Social Determinants of Health Data. TPJ 2018;22:18–078. 10.7812/TPP/18-078.

[17] Greenland S, Morgenstern H. Ecological Bias, Confounding, and Effect Modification. Int J Epidemiol 1989;18:269–74. 10.1093/ije/18.1.269.

[18] CDC/NCHS. About NHANES. National Health and Nutrition Examination Survey 2024. https://www.cdc.gov/nchs/nhanes/about/index.html (accessed March 18, 2026).

[19] CDC/NCHS. NHANES Questionnaires, Datasets, and Related Documentation n.d. https://wwwn.cdc.gov/nchs/nhanes/default.aspx (accessed May 27, 2026).

[20] Nguyen VK, Middleton LYM, Huang L, et al. Harmonized US National Health and Nutrition Examination Survey 1988-2018 for high throughput exposome-health discovery 2023. 10.1101/2023.02.06.23284573.

[21] Jaeger J. Digit Symbol Substitution Test: The Case for Sensitivity Over Specificity in Neuropsychological Testing. Journal of Clinical Psychopharmacology 2018;38:513. 10.1097/JCP.0000000000000941.

[22] CDC/NCHS. Cognitive Functioning (CFQ). National Health and Nutrition Examination Survey 1999-2000 Data Documentation, Codebook, and Frequencies 2005. https://wwwn.cdc.gov/Nchs/Data/Nhanes/Public/1999/DataFiles/CFQ.htm (accessed April 25, 2024).

[23] Middleton LYM, Walker E, Cockell S, et al. Exposome-wide association study of cognition among older adults in the National Health and Nutrition Examination Survey. Exposome 2025;5:osaf002. 10.1093/exposome/osaf002.

[24] Lee JY, Van Zandt S. Housing Tenure and Social Vulnerability to Disasters: A Review of the Evidence. Journal of Planning Literature 2019;34:156–70. 10.1177/0885412218812080.

[25] Ware LJ, Kim AW, Prioreschi A, et al. Social vulnerability, parity and food insecurity in urban South African young women: the healthy life trajectories initiative (HeLTI) study. J Public Health Policy 2021;42:373–89. 10.1057/s41271-021-00289-8.

[26] CDC/NCHS. Demographic Variables & Sample Weights (DEMO). National Health and Nutrition Examination Survey 1999-2000 Data Documentation, Codebook, and Frequencies 2002. https://wwwn.cdc.gov/Nchs/Data/Nhanes/Public/1999/DataFiles/DEMO.htm (accessed March 19, 2026).

[27] CDC/NCHS. Health Insurance (HIQ). National Health and Nutrition Examination Survey 1999-2000 Data Documentation, Codebook, and Frequencies 2004. https://wwwn.cdc.gov/Nchs/Data/Nhanes/Public/1999/DataFiles/HIQ.htm (accessed March 19, 2026).

[28] CDC/NCHS. Housing Characteristics (HOQ). National Health and Nutrition Examination Survey 1999-2000 Data Documentation, Codebook, and Frequencies 2004. https://wwwn.cdc.gov/Nchs/Data/Nhanes/Public/1999/DataFiles/HOQ.htm (accessed March 19, 2026).

[29] CDC/NCHS. Occupation (OCQ). National Health and Nutrition Examination Survey 1999-2000 Data Documentation, Codebook, and Frequencies 2008. https://wwwn.cdc.gov/Nchs/Data/Nhanes/Public/1999/DataFiles/OCQ.htm (accessed March 19, 2026).

[30] Fisher GG, Chaffee DS, Sonnega A. Retirement Timing: A Review and Recommendations for Future Research. WORKAR 2016;2:230–61. 10.1093/workar/waw001.

[31] CDC/NCHS. Food Security (FSQ). National Health and Nutrition Examination Survey 1999-2000 Data Documentation, Codebook, and Frequencies 2004. https://wwwn.cdc.gov/Nchs/Data/Nhanes/Public/1999/DataFiles/FSQ.htm (accessed March 19, 2026).

[32] Crane BM, Nichols E, Carlson MC, et al. Body Mass Index and Cognition: Associations Across Mid-to Late Life and Gender Differences. The Journals of Gerontology: Series A 2023;78:988–96. 10.1093/gerona/glad015.

[33] Topiwala A, Ebmeier KP. Effects of drinking on late-life brain and cognition. Evid Based Ment Health 2018;21:12–5. 10.1136/eb-2017-102820.

[34] Pampel FC, Krueger PM, Denney JT. Socioeconomic Disparities in Health Behaviors. Annu Rev Sociol 2010;36:349–70. 10.1146/annurev.soc.012809.102529.

[35] Schisterman EF, Cole SR, Platt RW. Overadjustment Bias and Unnecessary Adjustment in Epidemiologic Studies. Epidemiology 2009;20:488–95. 10.1097/EDE.0b013e3181a819a1.

[36] VanderWeele TJ. Principles of confounder selection. Eur J Epidemiol 2019;34:211–9. 10.1007/s10654-019-00494-6.

[37] Deguen S, Amuzu M, Simoncic V, et al. Exposome and Social Vulnerability: An Overview of the Literature Review. Int J Environ Res Public Health 2022;19:3534. 10.3390/ijerph19063534.

[38] R Core Team. R: A Language and Environment for Statistical Computing. Vienna, Austria: R Foundation for Statistical Computing; 2025.

[39] Lumley T. Analysis of Complex Survey Samples. J Stat Soft 2004;9. 10.18637/jss.v009.i08.

[40] Van Buuren S. Flexible Imputation of Missing Data, Second Edition. 2nd ed. Second Edition. | Boca Raton, Florida: CRC Press, [2019] |: Chapman and Hall/CRC; 2018. 10.1201/9780429492259.

[41] Buuren S van, Groothuis-Oudshoorn K. mice: Multivariate Imputation by Chained Equations in R. Journal of Statistical Software 2011;45:1–67. 10.18637/jss.v045.i03.

[42] Vassilaki M, Aakre JA, Castillo A, et al. Association of neighborhood socioeconomic disadvantage and cognitive impairment. Alzheimer’s &Dementia 2023;19:761–70. 10.1002/alz.12702.

[43] Kind AJH, Buckingham WR. Making Neighborhood-Disadvantage Metrics Accessible — The Neighborhood Atlas. N Engl J Med 2018;378:2456–8. 10.1056/NEJMp1802313.

[44] Hunt JFV, Vogt NM, Jonaitis EM, et al. Association of Neighborhood Context, Cognitive Decline, and Cortical Change in an Unimpaired Cohort. Neurology 2021;96:e2500–12. 10.1212/WNL.0000000000011918.

[45] Lamar M, Kershaw KN, Leurgans SE, et al. Neighborhood-level social vulnerability and individual-level cognitive and motor functioning over time in older non-Latino Black and Latino adults. Front Hum Neurosci 2023;17:1125906. 10.3389/fnhum.2023.1125906.

[46] Desai P, Bond J, Dhana A, et al. The Social Vulnerability Index and Incidence of Alzheimer Disease in a Population-Based Sample of Older Adults. Neurology 2025;104:e213464. 10.1212/WNL.0000000000213464.

[47] Andrew MK, Rockwood K. Social vulnerability predicts cognitive decline in a prospective cohort of older Canadians. Alzheimer’s & Dementia 2010;6:319–325.e1. 10.1016/j.jalz.2009.11.001.

[48] Armstrong JJ, Mitnitski A, Andrew MK, et al. Cumulative impact of health deficits, social vulnerabilities, and protective factors on cognitive dynamics in late life: a multistate modeling approach. Alz Res Therapy 2015;7:38. 10.1186/s13195-015-0120-7.

[49] Chen M, Tan X, Padman R. Social determinants of health in electronic health records and their impact on analysis and risk prediction: A systematic review. Journal of the American Medical Informatics Association 2020;27:1764–73. 10.1093/jamia/ocaa143.

[50] Fillit HM, Rockwood K, Young JB. Brocklehurst’s Textbook of Geriatric Medicine and Gerontology. 8. Aufl. S.l.: Elsevier Health Care - Lehrbücher; 2016.

