## Supplemental Tables and Figures for "Individual-level Social Vulnerability and Cognitive Function Among Older Adults in the United States National Health and Nutrition Examination Survey"

### SUPPLEMENTARY TABLES AND FIGURES

**Supplemental Table 1.** Summary of analytic variables with missing data imputed using Multiple Imputation by Chained Equations (MICE).

| Variable | n Missing | % Missing | Imputation method |
|---|---|---|---|
| **Social Vulnerability Index components** | | | |
| Poverty-Income Ratio | 633 | 10.6 | Predictive mean modeling |
| Education | 10 | 0.17 | Polytomous logistic regression |
| Health insurance | 59 | 0.99 | Polytomous logistic regression |
| Housing tenure | 61 | 1.02 | Polytomous logistic regression |
| Employment status | 6 | 0.10 | Polytomous logistic regression |
| Food security | 122 | 2.04 | Polytomous logistic regression |
| **Covariates** | | | |
| Smoking status | 9 | 0.15 | Polytomous logistic regression |
| Alcohol consumption | 1,386 | 23.14 | Predictive mean modeling |
| Body Mass Index (kg/m$^2$) | 460 | 7.68 | Predictive mean modeling |

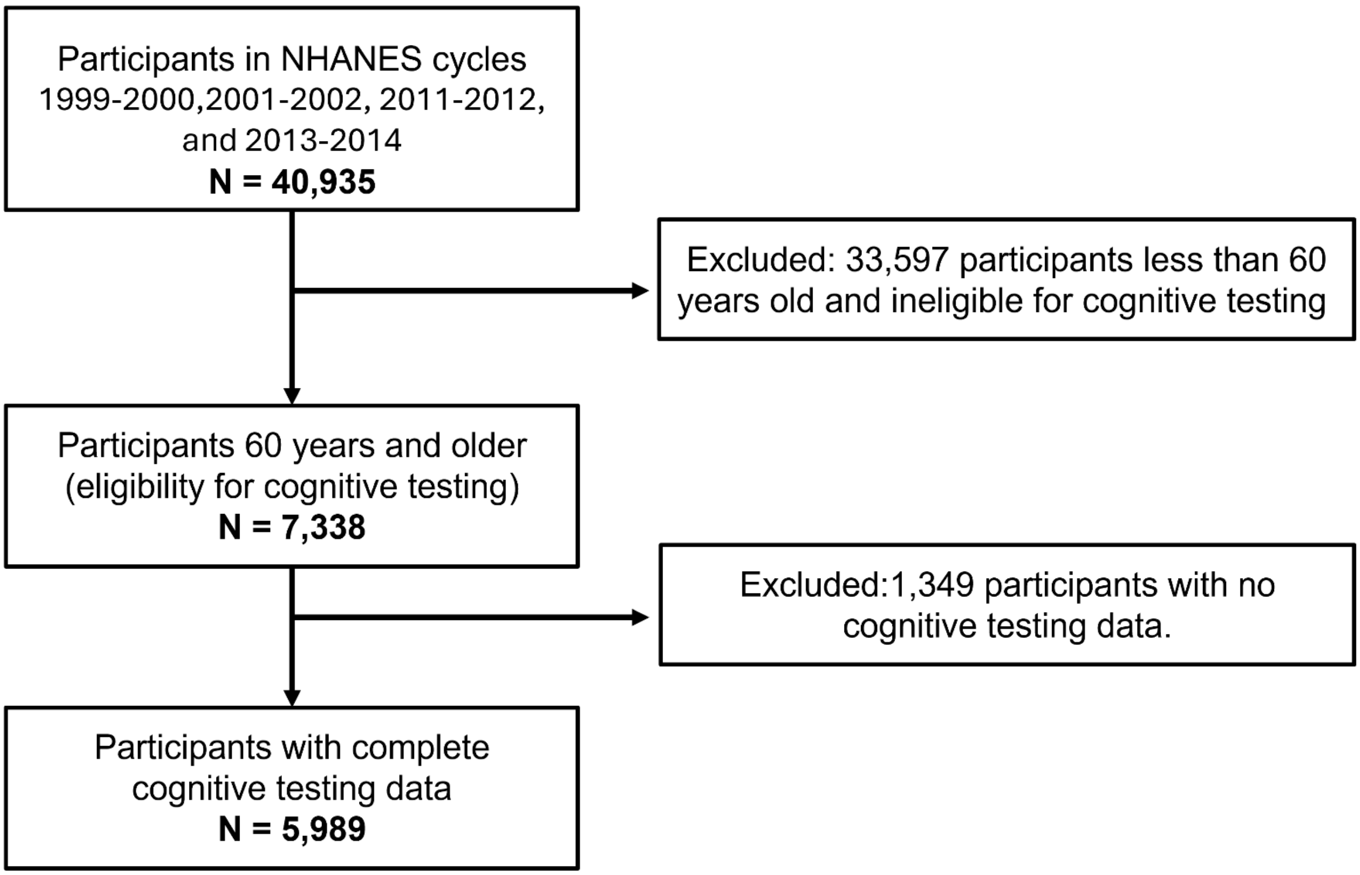


**Supplemental Figure 1.** Inclusion and exclusion flowcharts for the analytic sample in the National Health and Nutrition Examination Survey (NHANES), years 1999-2000,2001-2002, 2011-2012, and 2013-2014.

**Supplemental Table 2.** Comparative characteristics of excluded vs. included National Health and Nutrition Examination Survey participants (1999-2002 & 2011-2014 cycles), age ≥60 years old.

| Characteristic [a] | **Overall** (n=7,338) n (%) | **Excluded** (n= 1,349) n (%) | **Included** (n= 5,989) n (%) | *p-value* [b*] |
|---|---|---|---|---|
| **Social Vulnerability Index (iSVI),** mean (SD) | 1.46 (1.33) | 2.20 (1.44) | 1.36 (1.28) | <0.001 |
| Missing | 1,009 | 292 | 717 | |
| **iSVI Categories** | | | | <0.001 |
| Low | 3,981 (76.14) | 440 (53.65) | 3,541 (79.13) | |
| High | 2,348 (23.86) | 617 (46.35) | 1,731 (20.87) | |
| Missing | 1,009 | 292 | 717 | |
| **Poverty-Income Ratio** | | | | <0.001 |
| >1.5 | 4,003 (73.41) | 514 (56.15) | 3,489 (75.74) | |
| ≤1.5 | 2,438 (26.59) | 571 (43.85) | 1,867 (24.26) | |
| Missing | 897 | 264 | 633 | |
| **Education** | | | | <0.001 |
| Completed high school or above | 4,573 (75.49) | 558 (55.02) | 4,015 (78.52) | |
| Did not complete high school | 2,725 (24.51) | 761 (44.98) | 1,964 (21.48) | |
| Missing | 40 | 30 | 10 | |
| **Health Insurance** | | | | <0.001 |
| Private insurance | 3,633 (60.23) | 482 (45.26) | 3,151 (62.41) | |
| Government insurance | 3,060 (34.85) | 699 (49.60) | 2,361 (32.71) | |
| No insurance | 530 (4.91) | 112 (5.14) | 418 (4.88) | |
| Missing | 115 | 56 | 59 | |
| **Housing Tenure** | | | | <0.001 |
| Homeowner | 5,382 (81.70) | 859 (72.33) | 4,523 (83.06) | |
| Renter or Other | 1,842 (18.30) | 437 (27.67) | 1,405 (16.94) | |
| Missing | 114 | 53 | 61 | |
| **Employment Status** | | | | <0.001 |
| Employed | 1,588 (25.99) | 156 (12.99) | 1,432 (27.93) | |
| Retired | 4,393 (59.58) | 776 (60.23) | 3,617 (59.48) | |
| Unemployed | 1,341 (14.43) | 407 (26.78) | 934 (12.59) | |
| Missing | 16 | 10 | 6 | |
| **Food Security** | | | | <0.001 |
| Full food security | 5,910 (88.93) | 986 (83.98) | 4,924 (89.64) | |
| Less than full food security | 1,239 (11.07) | 296 (16.02) | 943 (10.36) | |
| Missing | 189 | 67 | 122 | |
| **Digit Symbol Substitution Test (DSST),** mean (SD) | 49.61 (17.73) | --- | 49.61 (17.73) | |
| Missing | 1,349 | 1,349 | 0 | |
| **Low Cognitive Performance** | | | | |
| No | 3,543 (73.58) | --- | 3,543 (73.58) | |
| Yes | 2,446 (26.42) | --- | 2,446 (26.42) | |
| Missing | 1,349 | 1,349 | 0 | |
| **Age at Baseline (years),** mean (SD) | 70.31 (7.25) | 73.79 (7.73) | 69.78 (7.03) | <0.001 |
| **Sex** | | | | 0.2 |
| Male | 3,549 (44.34) | 642 (41.97) | 2,907 (44.69) | |
| Female | 3,789 (55.66) | 707 (58.03) | 3,082 (55.31) | |
| **Race and Hispanic Origin** | | | | <0.001 |
| Non-Hispanic White | 3,762 (78.68) | 523 (63.16) | 3,239 (81.02) | |
| Mexican American & Other Hispanic | 1,595 (7.90) | 355 (13.07) | 1,240 (7.12) | |
| Non-Hispanic Black | 1,485 (8.72) | 341 (15.19) | 1,144 (7.74) | |
| Other Race | 496 (4.70) | 130 (8.58) | 366 (4.12) | |
| **NHANES Cycle Year** | | | | 0.002 |
| 1991-2000 | 1,834 (21.13) | 417 (26.82) | 1,417 (20.27) | |
| 2001-2002 | 1,872 (21.60) | 314 (20.15) | 1,558 (21.81) | |

| | Overall (n=7,338) | Excluded (n= 1,349) | Included (n= 5,989) | |
|---|---|---|---|---|
| 2011-2012 | 1,791 (27.77) | 369 (31.18) | 1,422 (27.25) | |
| 2013-2014 | 1,841 (29.51) | 249 (21.85) | 1,592 (30.67) | |
| **Smoking Status** | | | | 0.4 |
| Never smoker | 3,599 (48.93) | 707 (50.06) | 2,892 (48.77) | |
| Former smoker | 2,806 (39.33) | 460 (36.99) | 2,346 (39.68) | |
| Current smoker | 909 (11.73) | 167 (12.95) | 742 (11.55) | |
| Missing | 24 | 15 | 9 | |
| **Daily Alcohol Intake** | | | | 0.001 |
| <1 drink | 4,633 (87.27) | 542 (93.65) | 4,091 (86.78) | |
| ≥ 1 drink | 545 (12.73) | 33 (6.35) | 512 (13.22) | |
| Missing | 2,160 | 774 | 1,386 | |
| **Body Mass Index (kg/m$^2$),** mean (SD) | 28.70 (6.02) | 28.47 (6.39) | 28.73 (5.98) | 0.2 |
| Missing | 948 | 488 | 460 | |

*Note.* iSVI categories were derived from survey-weighted quartiles of iSVI, grouped as Low (quartile 1-3: 0 – 2; 79.1%) and High (quartile 4: 2.5 – 6; 20.9%), representing increasing levels of social vulnerability. Rows highlighted in grey are components of the iSVI. Low Cognitive Performance was defined as a DSST score in the lowest quartile (Q1; DSST ≤ 38). Abbreviations: SD, standard deviation.

[a] Mean (SD); n (%). Survey weighted means and standard errors were calculated for continuous variables. Unweighted frequencies and survey weighted percentages were calculated for categorical variables.

[b] Design-based Kruskal Wallis test; Pearson's $X^2$: Rao & Scott adjustment.

[*] $p<0.05$

**Supplemental Table 3.** Weighted baseline distribution of iSVI components among participants aged 60 years or older in the NHANES overall and stratified by iSVI categories.

| | **Overall** (n=5,989) | **Low iSVI** (n=3,541) | **High iSVI** (n=1,731) | |
|---|---|---|---|---|
| **Characteristic** [a] | **n (%)** | **n (%)** | **n (%)** | ***p-value*** [b*] |
| **Social Vulnerability Index (iSVI),** mean (SD) | 1.36 (1.28) | 0.80 (0.62) | 3.46 (0.85) | <0.001 |
| Missing | 717 | | | |
| **Poverty-Income Ratio** | | | | <0.001 |
| >1.5 | 3,489 (75.7) | 3,143 (91.9) | 278 (15.5) | |
| ≤1.5 | 1,867 (24.2) | 398 (8.4) | 1,453 (84.5) | |
| Missing | 633 | | | |
| **Education** | | | | <0.001 |
| Completed high school or above | 4,015 (78.5) | 3,033 (90.1) | 554 (38.4) | |
| Did not complete high school | 1,964 (21.5) | 508 (9.9) | 1,177 (61.6) | |
| Missing | 10 | | | |
| **Health Insurance** | | | | <0.001 |
| Private insurance | 3,151 (62.4) | 2,438 (72.5) | 377 (24.9) | |
| Government insurance | 2,361 (32.7) | 1,002 (24.8) | 1,088 (61.6) | |
| No insurance | 418 (4.8) | 101 (2.7) | 266 (13.5) | |
| Missing | 59 | | | |
| **Housing Tenure** | | | | <0.001 |
| Homeowner | 4,523 (83.1) | 3,222 (92.8) | 793 (46.5) | |
| Renter or Other | 1,405 (16.9) | 319 (7.3) | 938 (53.5) | |
| Missing | 61 | | | |
| **Employment Status** | | | | <0.001 |
| Employed | 1,432 (27.9) | 1,076 (33.6) | 192 (9.5) | |
| Retired | 3,617 (59.5) | 2,229 (59.1) | 957 (58.7) | |
| Unemployed | 934 (12.6) | 236 (7.4) | 582 (31.8) | |
| Missing | 6 | | | |
| **Food Security** | | | | <0.001 |
| Full food security | 4,924 (89.6) | 3,429 (97.3) | 972 (58.5) | |
| Less than full food security | 943 (10.4) | 112 (2.5) | 759 (41.5) | |
| Missing | 122 | | | |

*Note.* iSVI categories were derived from survey-weighted quartiles of iSVI, grouped as Low (quartile 1-3: 0 – 2; 79.1%) and High (quartile 4: 2.5 – 6; 20.9%), representing increasing levels of social vulnerability. Abbreviations: SD, standard deviation.

[a] Mean (SD); n (%). Survey weighted means and standard errors were calculated for continuous variables. Unweighted frequencies and survey weighted percentages were calculated for categorical variables.

[b] Design-based Kruskal Wallis test; Pearson's $X^2$: Rao & Scott adjustment.

[*] $p<0.05$

**Supplemental Figure 2.** Histogram of Digit Symbol Substitution Test (DSST) Scores with Threshold for Low Cognitive Performance.

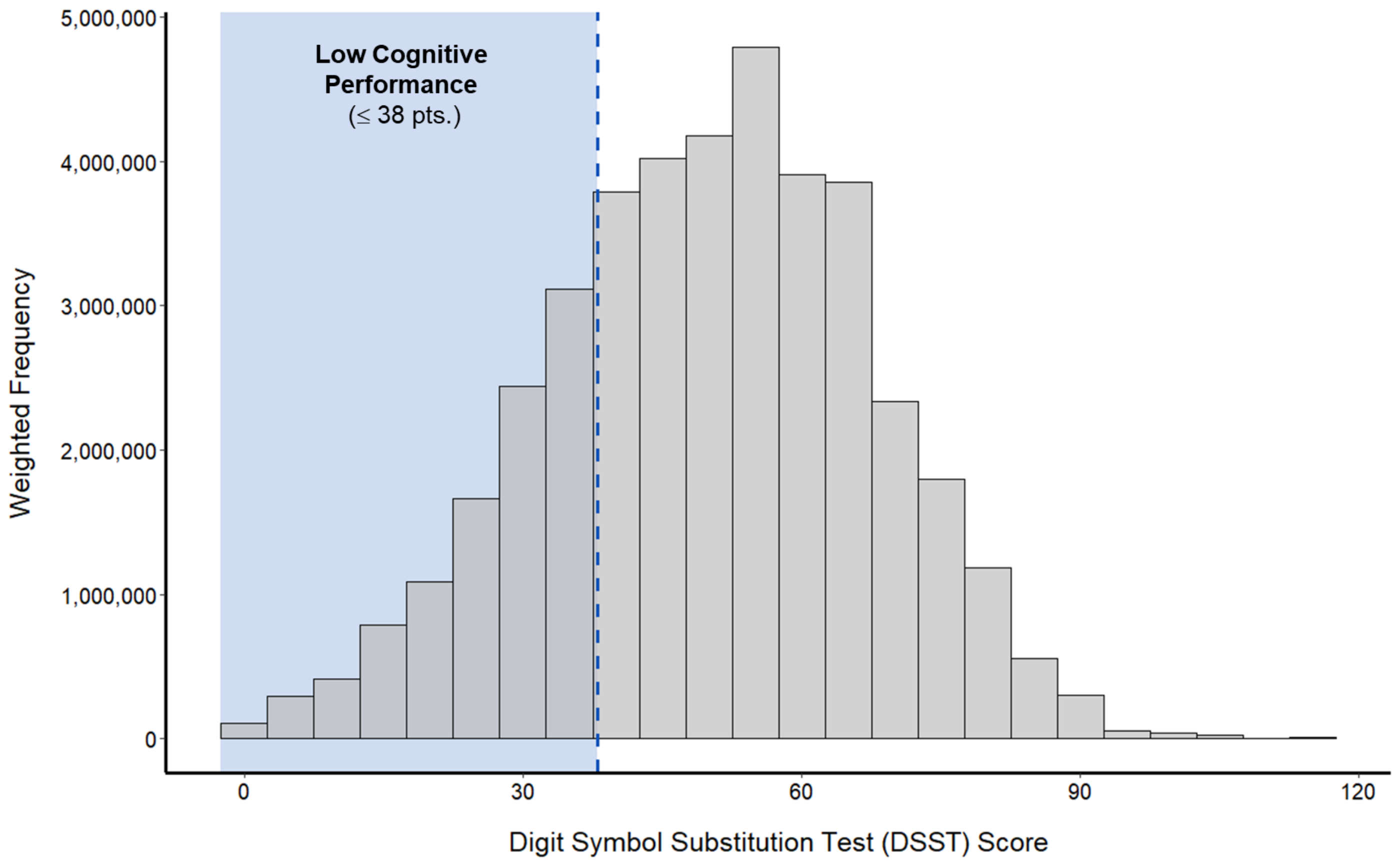


[a] Histogram showing the distribution of DSST scores among participants aged ≥60 years. The vertical line dictates the survey-weighted 25th percentile cutoff (DSST ≤38). Scores at or below this cutoff were classified as low cognitive performance, and scores in quartiles 2-4 (DSST >38) were classified as normal cognitive performance for comparison in supplemental descriptive analyses.

**Supplemental Table 4.** Weighted baseline characteristics of included continuous National Health and Nutrition Examination Survey (NHANES, N = 5,989) participants age ≥60 years old, overall and stratified by low cognitive performance status.

| | **Overall** (n=5,989) | **Low Cognitive Performance** (n=2,446) | **Normal Cognitive Performance** (n=3,543) | |
|---|---|---|---|---|
| **Characteristic** [a] | **n (%)** | **n (%)** | **n (%)** | ***p*** [b*] |
| **Social Vulnerability Index (iSVI),** mean (SD) | 1.36 (1.28) | 2.26 (1.42) | 1.04 (1.05) | <0.001 |
| Missing | 717 | 319 | 398 | |
| **iSVI Categories** | | | | <0.001 |
| Low | 3,541 (79.13) | 965 (53.45) | 2,576 (88.16) | |
| High | 1,731 (20.87) | 1,162 (46.55) | 569 (11.84) | |
| Missing | 717 | 319 | 398 | |
| **Poverty-Income Ratio** | | | | <0.001 |
| >1.5 | 3,489 (75.74) | 1,015 (52.46) | 2,474 (83.99) | |
| ≤1.5 | 1,867 (24.26) | 1,153 (47.54) | 714 (16.01) | |
| Missing | 633 | 278 | 355 | |
| **Education** | | | | <0.001 |
| Completed high school or above | 4,015 (78.52) | 1,030 (51.20) | 2,985 (88.29) | |
| Did not complete high school | 1,964 (21.48) | 1,408 (48.80) | 556 (11.71) | |
| Missing | 10 | 8 | 2 | |
| **Health Insurance** | | | | <0.001 |
| Private insurance | 3,151 (62.41) | 958 (45.07) | 2,193 (68.59) | |
| Government insurance | 2,361 (32.71) | 1,228 (48.66) | 1,133 (27.03) | |
| No insurance | 418 (4.88) | 230 (6.28) | 188 (4.39) | |
| Missing | 59 | 30 | 29 | |
| **Housing Tenure** | | | | <0.001 |
| Homeowner | 4,523 (83.06) | 1,622 (71.15) | 2,901 (87.33) | |
| Renter or Other | 1,405 (16.94) | 798 (28.85) | 607 (12.67) | |
| Missing | 61 | 26 | 35 | |
| **Employment Status** | | | | <0.001 |
| Employed | 1,432 (27.93) | 372 (12.74) | 1,060 (33.37) | |
| Retired | 3,617 (59.48) | 1,564 (69.09) | 2,053 (56.04) | |
| Unemployed | 934 (12.59) | 505 (18.18) | 429 (10.58) | |
| Missing | 6 | 5 | 1 | |
| **Food Security** | | | | <0.001 |
| Full food security | 4,924 (89.64) | 1,818 (80.76) | 3,106 (92.81) | |
| Less than full food security | 943 (10.36) | 574 (19.24) | 369 (7.19) | |
| Missing | 122 | 54 | 68 | |
| **Digit Symbol Substitution Test (DSST),** mean (SD) | 49.61 (17.73) | 26.99 (8.88) | 57.73 (12.22) | <0.001 |
| **Age (years),** mean (SD) | 69.78 (7.03) | 73.23 (7.28) | 68.54 (6.50) | <0.001 |
| **Sex** | | | | 0.2 |
| Male | 2,907 (44.69) | 1,295 (46.44) | 1,612 (44.07) | |
| Female | 3,082 (55.31) | 1,151 (53.56) | 1,931 (55.93) | |
| **Race and Hispanic Origin** | | | | <0.001 |
| Non-Hispanic White | 3,239 (81.02) | 971 (64.84) | 2,268 (86.83) | |
| Mexican American & Other Hispanic | 1,240 (7.12) | 743 (15.58) | 497 (4.09) | |
| Non-Hispanic Black | 1,144 (7.74) | 632 (15.94) | 512 (4.79) | |
| Other Race | 366 (4.12) | 100 (3.64) | 266 (4.29) | |
| **NHANES Cycle Year** | | | | <0.001 |
| 1991-2000 | 1,417 (20.27) | 706 (27.93) | 711 (17.52) | |
| 2001-2002 | 1,558 (21.81) | 680 (26.31) | 878 (20.20) | |

| | Overall | Low Cognitive Performance | Normal Cognitive Performance | |
|---|---|---|---|---|
| | (n=5,989) | (n=2,446) | (n=3,543) | |
| **Characteristic** [a] | **n (%)** | **n (%)** | **n (%)** | ***p*** [b*] |
| 2011-2012 | 1,422 (27.25) | 527 (21.00) | 895 (29.49) | |
| 2013-2014 | 1,592 (30.67) | 533 (24.75) | 1,059 (32.79) | |
| **Smoking Status** | | | | 0.10 |
| Never smoker | 2,892 (48.77) | 1,171 (48.29) | 1,721 (48.94) | |
| Former smoker | 2,346 (39.68) | 924 (38.33) | 1,422 (40.17) | |
| Current smoker | 742 (11.55) | 346 (13.38) | 396 (10.89) | |
| Missing | 9 | 5 | 4 | |
| **Daily Alcohol Intake** | | | | <0.001 |
| <1 drink | 4,091 (86.78) | 1,553 (91.63) | 2,538 (85.31) | |
| ≥ 1 drink | 512 (13.22) | 150 (8.37) | 362 (14.69) | |
| Missing | 1,386 | 743 | 643 | |
| **Body Mass Index (kg/m$^2$),** mean (SD) | 28.73 (5.98) | 28.35 (6.03) | 28.85 (5.96) | 0.019 |
| Missing | 460 | 288 | 175 | |

*Note.* Low cognitive performance was defined as DSST scores in the lowest survey-weighted quartile (DSST ≤38). Participants with scores in quartiles 2-4 (DSST >38) were classified as normal cognitive performance for comparison. iSVI categories were derived from survey-weighted quartiles of iSVI, grouped as Low (quartile 1-3) and High (quartile 4), representing increasing levels of social vulnerability. Rows highlighted in grey are components of the iSVI. Abbreviations: SD, standard deviation.

[a] Mean (SD); n (%). Survey weighted means and standard errors were calculated for continuous variables. Unweighted frequencies and survey weighted percentages were calculated for categorical variables.

[b] Design-based Kruskal Wallis test; Pearson's $X^2$: Rao & Scott adjustment.

[*] $p<0.05$

**Supplemental Figure 3.** Distribution of individual-level Social Vulnerability Index (iSVI) scores with category cutoffs.

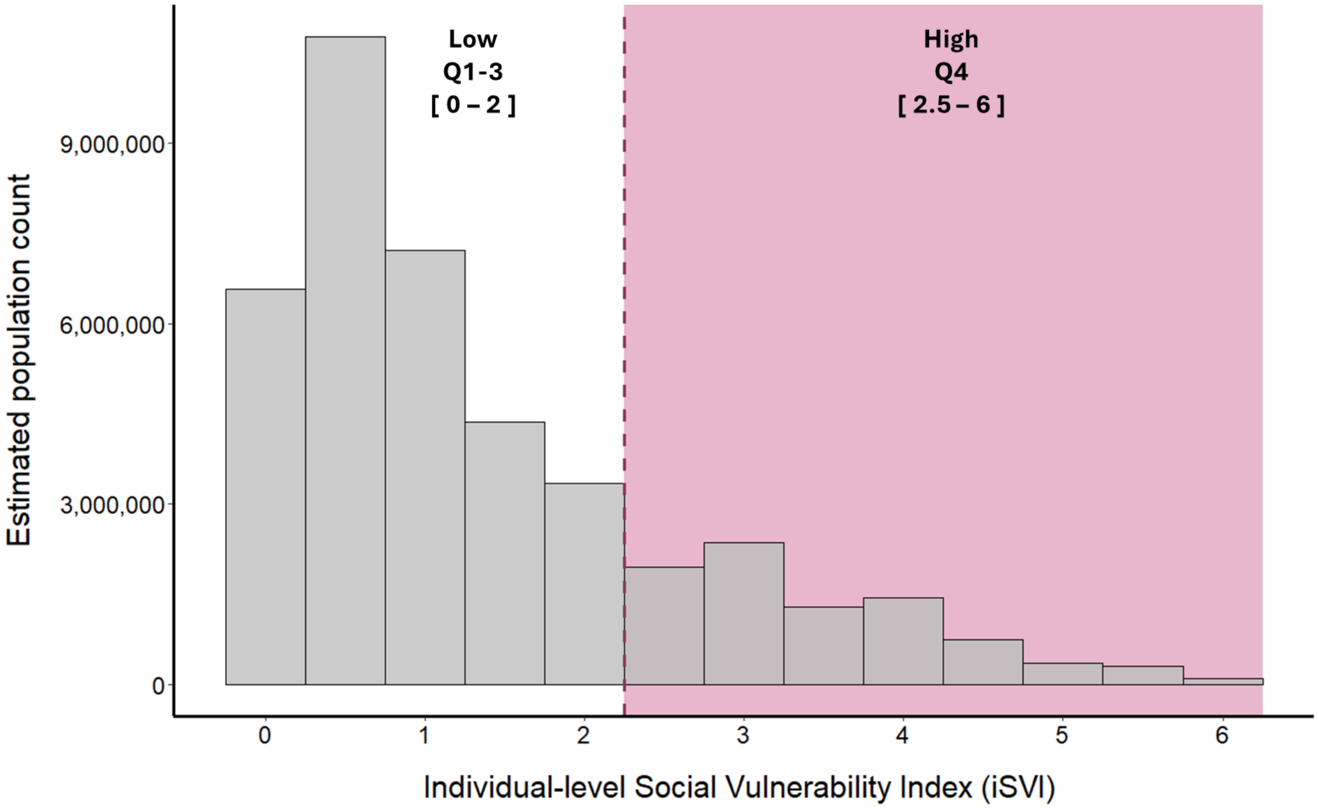


[a] Histogram showing the weighted distribution of iSVI scores among study participants aged ≥60 years. The dashed vertical line marks the cutoff used to define iSVI categories: low (quartiles 1- 3; Q1-Q3 ) and high (quartile 4; Q4)

**Supplemental Table 5.** Effect modification of the association between iSVI and DSST scores among NHANES participants aged ≥60 years (N = 5,989).

| | Continuous of iSVI | | Categories of iSVI | | |
|---|---|---|---|---|---|
| Models | **Per one unit increase** β (95% CI) | ***P for interaction*** | **Low** | **High** β (95% CI) | ***P for interaction*** |
| **iSVI x Age [a]** | | | | | |
| < 70 years old | **-5.43** (-5.96, -4.89) | 0.59 | Ref | **-15.25** (-17.34, -13.16) | **0.02** |
| ≥ 70 years old | **-5.63** (-6.24, -5.02) | | Ref | **-12.49** (-14.04, -10.94) | |
| **iSVI x Sex [b]** | | | | | |
| Male | **-5.35** (-5.86, -4.83) | 0.83 | Ref | **-14.08** (-15.52, 12.63) | 0.35 |
| Female | **-5.27** (-5.83, -4.71) | | Ref | **-13.15** (-14.93, 11.36) | |
| **iSVI x Race and Hispanic Origin [c]** | | | | | |
| Non-Hispanic White | **-5.52** (-6.04, -5.01) | 0.61 | Ref | **-14.87** (-16.68, -13.05) | 0.16 |
| Not Non-Hispanic White | **-5.38** (-5.83, -4.92) | | Ref | **-13.45** (-14.99, -11.91) | |

*Note.* iSVI categories were derived from survey-weighted quartiles of iSVI, grouped as Low (quartile 1-3) and High (quartile 4), representing increasing levels of social vulnerability. Abbreviations: SD, standard deviation. Abbreviations: CI, Confidence interval; iSVI, individual-level Social Vulnerability Index; DSST, Digit Symbol Substitution Test; NHANES, National Health and Nutrition Examination Survey; Ref, Reference group.

[a] Adjusted for age group (<70 vs. ≥70 years), sex, race and Hispanic origin, NHANES cycle, and a multiplicative interaction term between iSVI and age group.

[b] Adjusted for age, sex, race and Hispanic origin, NHANES cycle, and a multiplicative interaction term between iSVI and sex.

[c] Adjusted for age, sex, race/ethnicity category (non-Hispanic White vs. not non-Hispanic White), NHANES cycle, and a multiplicative interaction term between iSVI and race/ethnicity category.

[*] $p<0.05$
